# Polygenic Background and Penetrance of Pathogenic Variants in Hypertrophic and Dilated Cardiomyopathies

**DOI:** 10.1101/2025.06.20.25329138

**Authors:** Sarah A. Abramowitz, Lily Hoffman-Andrews, David Zhang, Renae Judy, Thomas P. Cappola, Sharlene M. Day, Nosheen Reza, Anjali T. Owens, Scott M. Damrauer, Michael G. Levin

## Abstract

**Importance:** Polygenic background modifies variant penetrance in hypertrophic (HCM) and dilated (DCM) cardiomyopathy, diseases with opposing morphologic characteristics and inversely related genetic pathways. Whether polygenic susceptibility for one disease protects against monogenic risk for the other remains unexplored.

****Objective**:** To characterize if polygenic background bidirectionally modifies pathogenicity of established rare variants associated with HCM and DCM.

****Design**:** Cross-sectional study.

****Setting**:** The Penn Medicine BioBank (PMBB).

****Participants**:** Volunteers enrolled in PMBB with available electronic health record and genotyping data.

****Exposures**:** Normalized polygenic scores (PGS) for HCM and DCM, as well as carrier status of pathogenic variants in established HCM or DCM genes.

****Main Outcomes**:** HCM and DCM defined using electronic health record diagnosis and procedure code, as well as echocardiogram measurements derived from medical records.

****Results**:** This study included 49,434 PMBB participants. An increased HCM PGS was associated with significantly increased left ventricular ejection fraction (LVEF), decreased left ventricular internal diameter at end-diastole (LVIDd), and increased interventricular septal thickness (IVS) (p<0.001). An increased DCM PGS was significantly (p<0.001) associated with decreased LVEF and increased LVIDd, but was not associated with IVS. A one standard deviation increase in HCM PGS was associated with increased risk of HCM (OR 1.8; 95% CI 1.6-2.0; p=9.6x10^-25^) and decreased risk of DCM (OR 0.69; 95% CI 0.64-0.74; p=4.3x10^-22^). A one standard deviation increase in DCM PGS was associated with an increased risk of DCM (OR 1.6; 95% CI 1.5-1.7; p=1.7x10^-40^) and decreased risk of HCM (OR 0.69; 95% CI 0.63-0.76; p=3.0x10^-13^). Monogenic and polygenic risk terms had significant, independent effects when combined in models of disease status and echocardiographic measurements; the additional inclusion of either an HCM or DCM PGS improved the discrimination of models of HCM and DCM that included age, sex, and monogenic variant status (>95% probability of difference in AUROC).

**Conclusions and Relevance:** HCM and DCM risk are markedly modified by polygenic background which exists on an overlapping spectrum. Consideration of polygenic background may offer clinical value through improving understanding and prediction of these inherited cardiomyopathies.

**Key Points:** *Question:* How is risk for hypertrophic and dilated cardiomyopathy modified by polygenic background?

*Findings:* Polygenic scores for HCM and DCM associate with clinical and echocardiographic measures relevant to both diseases and inversely modify the penetrance of pathogenic variants.

*Meaning:* Polygenic background contributes to HCM and DCM susceptibility, and exists on an overlapping spectrum.

## Introduction

Hypertrophic (HCM) and dilated (DCM) cardiomyopathies are the most prevalent inherited forms of cardiomyopathy, affecting as many as one in 200 or 250 individuals respectively.^1,2^ HCM and DCM are characterized by contrasting cardiac morphology; HCM is characterized by left ventricular hypertrophy and hyperdynamic systolic function, whereas DCM features left ventricular dilation and systolic dysfunction. While these cardiomyopathies can be partially attributed to rare pathogenic variants in more than two dozen rigorously curated genes often affecting the cardiac sarcomere, HCM and DCM cannot be completely explained by rare genetic variation.^3–5^ Although HCM is classically considered a Mendelian condition, genetic testing for a causal variant is positive in around 25-35% of cases.^6–9^ Similarly, known pathogenic variants for dilated cardiomyopathy are identified in an estimated 15-30% of cases.^2,10,11^ Most carriers of pathogenic variants do not develop disease, reflecting variable penetrance.^1,10,12^

Recent population-scale genomic studies of HCM and DCM have identified a role for common genetic variants. A substantial portion of the heritability of HCM and DCM is attributable to the cumulative burden of these common variants with individually-small effect sizes.^13–15^ Polygenic scores (PGSs) for HCM and DCM, which aggregate genetic risk attributable to common genetic variants, have separately been shown to stratify disease risk among those with and without known monogenic variants for the corresponding disease.^13,14,16,17^

Building on prior studies that have identified several individual common variants that are inversely associated with both HCM and DCM,^14,15,18,19^ we hypothesized that PGSs for HCM and DCM would have bidirectional associations with clinical and echocardiographic outcomes. To test this hypothesis, we assessed the contributions of monogenic variants and PGSs to HCM and DCM and their associated echocardiographic traits, and evaluated whether PGS for HCM and DCM bidirectionally modify clinical and echocardiographic penetrance of pathogenic variants.

## Methods

### Study Population

The Penn Medicine BioBank (PMBB) is an electronic health record (EHR)-linked biobank comprised of consenting individuals receiving care within the Penn Medicine health system. PMBB details have been previously described.^20^ The current analysis considered individuals enrolled between 2008 and 2022, with health records available as early as 1994 through 2024. Analysis was performed in June 2025.

### Echocardiographic Outcomes

Clinically obtained echocardiography measurements were extracted from PMBB participant medical records. Measurements of left ventricular ejection fraction (LVEF), left ventricular internal diameter at end-diastole (LVIDd), and interventricular septal thickness (IVS) were compiled. For a given measurement, physiologically implausible values were removed (ie. LVEF < 0%). Values exceeding a median absolute deviation threshold of five (calculated using the ‘Routliers’ R package) were also excluded. Each individual’s maximum LVIDd and minimum LVEF were considered for evaluations of DCM, and maximum LVEF and maximum IVS were considered for evaluations of HCM.

### Clinical Outcomes

Designations of HCM/DCM case and non-case status were made using electronic health record diagnosis codes from the 9^th^ and 10^th^ revisions of the *International Classification of Diseases* (ICD) as well as Current Procedural Terminology (CPT) codes. A table listing these diagnosis codes can be found in eTable 1. Individuals with at least one diagnosis code for HCM phenocopies including amyloidosis and metabolic disorders were excluded (eTable 1). DCM cases were defined by the presence of a DCM code in the absence of an antecedent coronary artery disease (CAD) code, as previously described.^21,22^ HCM cases had at least two HCM codes from different dates. Individuals with both HCM and DCM codes were excluded. Non-cases were defined by the absence of DCM or HCM case status, and the absence of any heart failure code. Clinical outcome definitions were validated with the previously described echocardiogram data, considering measurement-cutoff based diagnostic criteria for these conditions (eMethods).^22,23^

### Rare Variant Classification

ClinGen is an NIH-funded collaborative resource that organizes expert panels to curate the strength of evidence supporting clinically-relevant gene-disease associations.^24^ Among carriers of rare variants in genes designated by ClinGen to have strong or definitive evidence for causing HCM and DCM, pathogenicity was determined by a genetic counselor (LH-A) in accordance with American College of Medical Genetics guidelines.^25^ Strong/definitive evidence HCM genes were *ACTN2, ACTC1, ALPK3, CSRP3, FHOD3, MYBPC3, MYH7, MYL2, MYL3, PLN, TNNC1, TNNI3, TNNT2*, and *TPM1*.^5^ Strong/definitive evidence DCM genes were *BAG3, DES, DSP, FLNC, LMNA, MYH7, PLN, RBM20, SCN5A, TNNC1, TNNT2,* and *TTN*.^4^ Loss of function variants in high percentage spliced in exons (>90%) of *TTN* were additionally considered pathogenic for DCM.^26^

### Polygenic Scoring

We used a published HCM PGS (PGS Catalog ID PGS004910) generated with *PRS-CS* from a genome-wide association study (GWAS) of 5,900 individuals with HCM and 68,359 without.^16,27^ We applied the same method (*PRS-CS-auto*) to create a DCM PGS using summary data from a GWAS of 9,365 individuals with DCM and 946,368 without (eMethods).^13^ Neither the HCM nor DCM summary data includes PMBB participants.

PGSs for each PMBB participant were calculated with *pgsc_calc* using the described PGS weights and imputed genotypes.^28^ Because PGS distributions may be influenced by allele frequency differences related to genetic ancestry, we applied the Z_norm2 principal component-based method for normalizing score mean and variance across populations to a standard normal distribution using external 1000 Genomes Project and Human Genomes Diversity Project reference data.^28^

### Statistical Analysis

Statistical analysis was performed using R version 4.3.2 (R Foundation for Statistical Computing, Vienna, Austria). The cross-trait genetic correlation between HCM and DCM was estimated using LD-score regression.^29^ Correlation of individual-level HCM and DCM scores was assessed with Pearson correlation. Pooled penetrance of pathogenic HCM and DCM variants was estimated by dividing the number of affected pathogenic variant carriers by the total number of pathogenic variant carriers for each condition and implementing binomial exact confidence intervals. Multivariable logistic regression was used to evaluate the risk of prevalent HCM and DCM associated with polygenic risk scores and/or pathogenic variant status, adjusting for age at PMBB enrollment and sex; genetic principal components were not included as the mean and variance of each score were normalized using genetic principal components.^28^ Marginal predictions were generated and plotted with the *‘marginaleffects*’ R package to visualize the relationship between polygenic and monogenic risk. Calibration and discrimination of these models were respectively assessed with Brier scores and area under the receiver operator characteristic curve (AUROC), which were calculated with resampling using V-fold cross validation (7 repeats of 10 folds). We assessed the probability of an incremental difference in Brier score and AUROC between base predictive models that included age and sex, and those that included combinations of polygenic and monogenic risk factors using the *‘tidyposterior’* R package, deeming a probability of <95% as statistically equivalent. We tested associations between polygenic scores and echocardiogram measurements using linear regression, adjusted for sex and age at which a given measurement was taken. To account for multiple comparisons (two PGS tested for two primary clinical outcomes [HCM and DCM] totaling four tests), we applied a Bonferroni correction, adjusting our significance threshold to p<0.0125.

We conducted our primary analyses in the full PMBB cohort using PCA-normalized PGSs as described above. As a sensitivity analysis, this approach was replicated in subsets of the PMBB population genetically similar to European and African reference populations (eMethods).

## Results

### Clinical and Echocardiographic Characteristics of HCM & DCM

Among 49,434 unrelated participants of the Penn Medicine BioBank with both medical records and genetic data, we identified 419 HCM cases, 895 DCM cases, and 39,421 non-cases (Figure 1). 21,674 participants had at least one relevant echocardiogram measure; those with measurements had a median of two recorded echocardiograms (interquartile range 4). Participant demographics are available in eTables 2-3.

**Figure 1:**
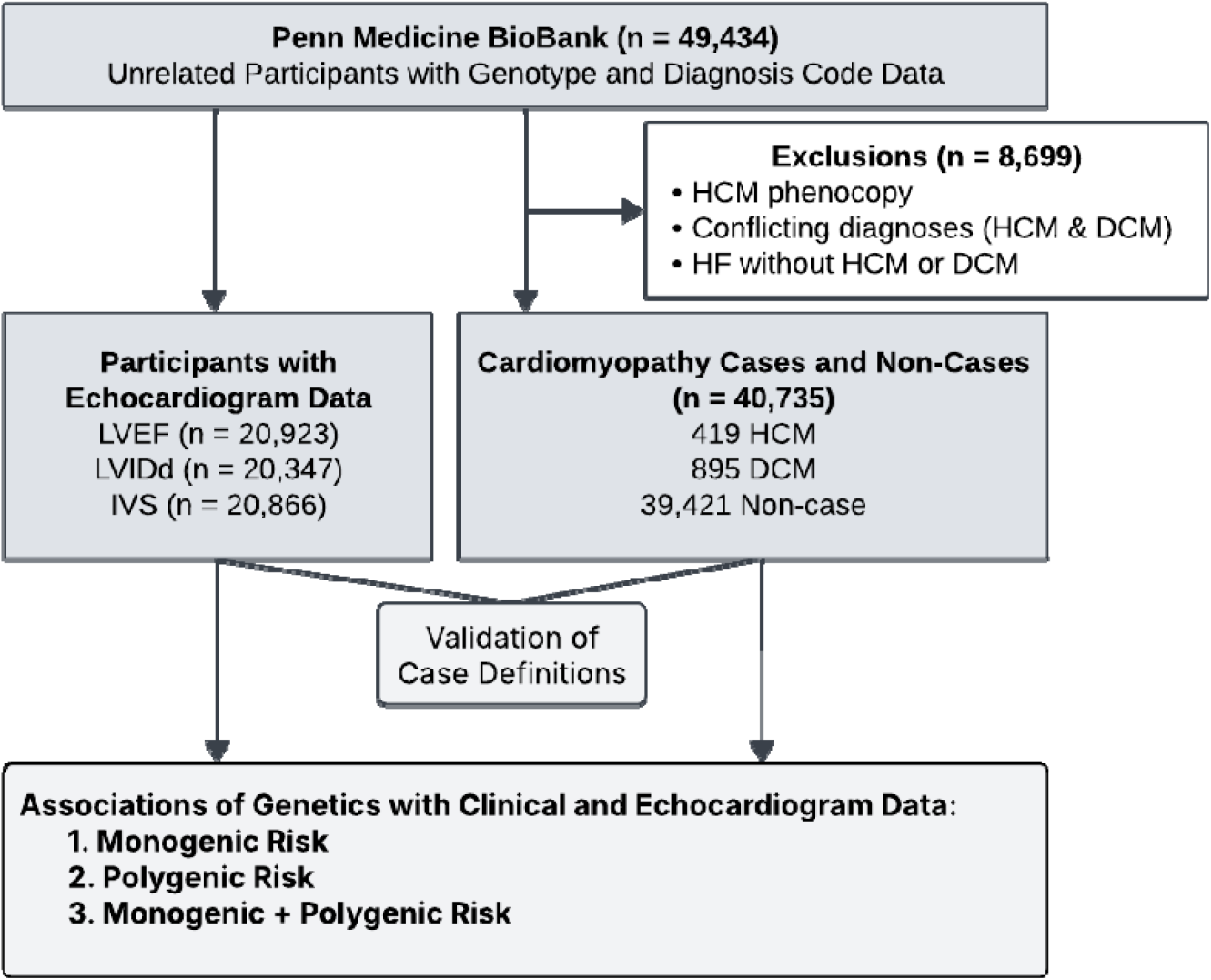
Sample filtering and workflow. From an original sample of unrelated participants, the subset of patients with echocardiogram data was identified. Individuals were separately classified for HCM or DCM case and non-case status, excluding individuals with conflicting diagnosis codes, heart failure, or an HCM phenocopy such as amyloidosis. After validating case definitions with echocardiogram data, associations of genetics with clinical and echocardiogram data were separately tested.

Cardiomyopathy definitions were validated by testing for associations between cardiomyopathy phenotypes and echocardiographic measures consistent with HCM and DCM in the subset of individuals with available echocardiographic data. Echocardiographic measurements were available from as early as 2009, through 2024. Individuals with HCM had significantly increased IVS (median 1.6 cm vs 1.1 cm; p = 6.17x10^-140^) and LVEF (median 75% vs 65%; p = 2.5x10^-74^) compared with non-cases. Individuals with DCM had significantly increased LVIDd (median 5.9 cm vs 4.7 cm; p = 1.1x10^-277^) and significantly decreased LVEF (median 30% vs 60%; p < 2.2x10^-308^) compared with non-cases. (eFigure 1). Our results internally validate our diagnosis code-derived clinical outcome definitions (eResults).

### Disease Associations of Pathogenic Variants

We first assessed the relationships between monogenic risk (characterized by the presence or absence of a pathogenic variant in a gene definitively associated with HCM or DCM) and prevalent diagnoses of HCM and DCM. In the 40,735 case/non-case participants, 259 had a pathogenic variant in an HCM gene, and 488 had a pathogenic variant in a DCM gene (eTable 4). 99 participants carried pathogenic variants in genes associated with both HCM and DCM (*TNNT2*, *MYH7, PLN, TNNC1*). Of 419 HCM cases, 84 (20%) were found to have a pathogenic variant (eFigure 2); pathogenic HCM variant status conferred a 65-fold increase in HCM risk (95% CI 48-86; p=8.0x10^-175^). Of 895 individuals with DCM, 94 (11%) carried pathogenic DCM variants (eFigure 2); pathogenic DCM variant status conferred a 13-fold risk of DCM (95% CI 10-17; p=1.2x10^-96^). The estimated pooled penetrance was 33% (95% CI 27-39%) for pathogenic HCM variants, and 21% (95% CI 18-25%) for pathogenic DCM variants. These results demonstrate that although pathogenic variants in HCM and DCM are not fully penetrant, they are associated with substantial increases in disease risk.

**Figure 2:**
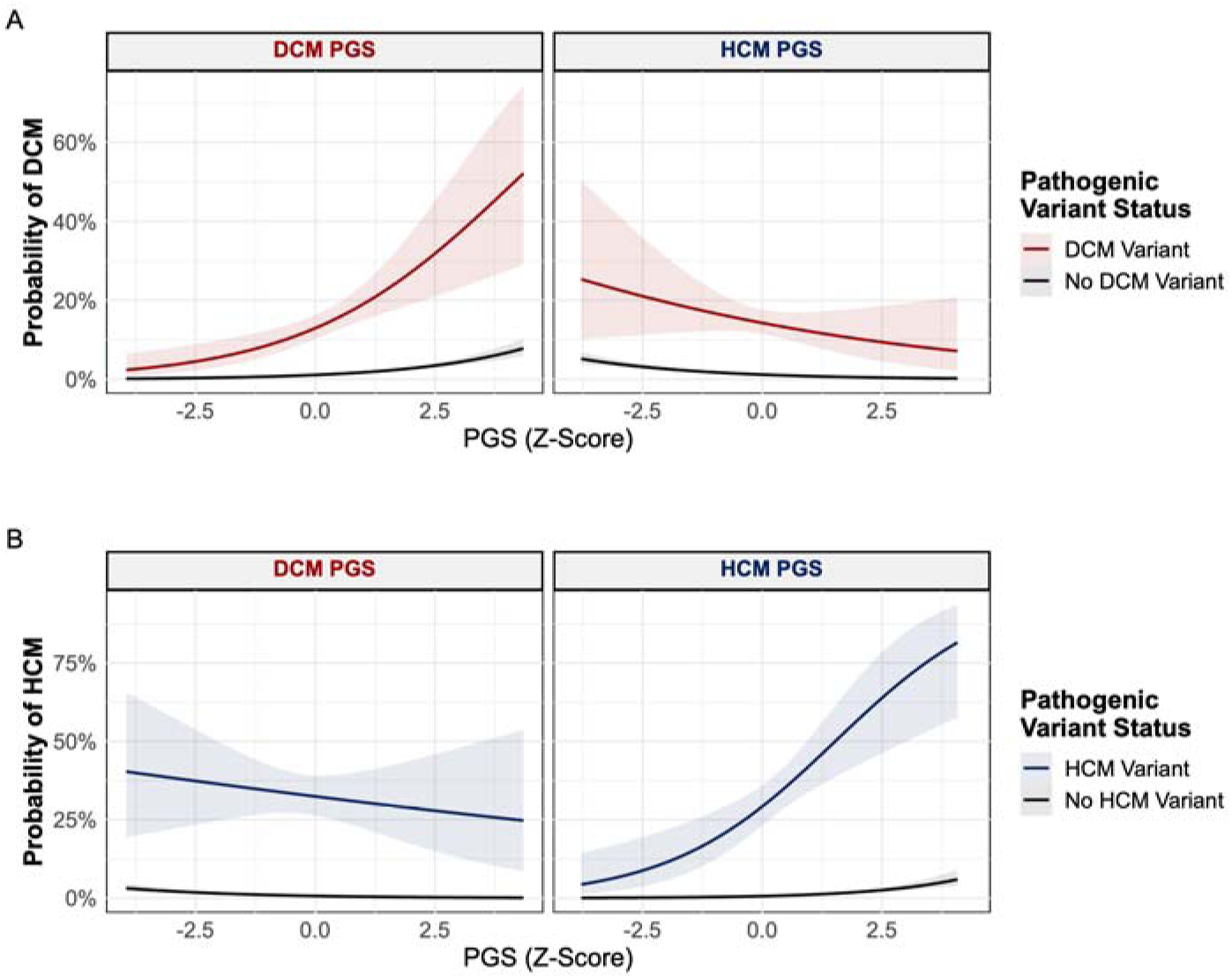
Effects of polygenic and monogenic risk on HCM and DCM probability. Logistic regression models including include polygenic risk, monogenic risk, a polygenic:monogenic interaction term, sex, and age at time of biobank enrollment were used for marginal prediction of clinical cardiomyopathy outcomes. **A)** Prediction of DCM in carriers of a DCM pathogenic variant, using a DCM (left) and HCM (right) PGS. **B)** Prediction of HCM in carriers of a HCM pathogenic variant, using a DCM (left) and HCM (right) PGS.

### Associations of Cardiomyopathy Polygenic Scores with Disease

Because HCM and DCM have been reported to exist along a phenotypic spectrum, and individual loci have been reported to be shared between the two diseases, we sought to quantify the similarity in their genetic architecture. At the summary-level, the cross-trait genetic correlation was -0.53 (p=5.10x10^-10^), and at the individual level there was modest negative correlation between DCM and HCM polygenic scores (r = -0.24; p<2.2x10^-16^).

Next, we sought to evaluate associations between PGS for HCM and DCM with disease status in PMBB, to determine whether each PGS demonstrated bidirectional associations with HCM and DCM. A one standard deviation increase in HCM PGS was associated with increased risk of HCM (OR 1.8; 95% CI 1.6-2.0; p=9.6x10^-25^) and decreased risk of DCM (OR 0.69; 95% CI 0.64-0.74; p=4.3x10^-22^). A one standard deviation increase in DCM PGS was associated with an increased risk of DCM (OR 1.6; 95% CI 1.5-1.7; p=1.7x10^-40^) and decreased risk of HCM (OR 0.69; 95% CI 0.63-0.76; p=3.0x10^-13^). Risk distributions of the DCM and HCM PGS among individuals with/without DCM and HCM are presented in eFigure 3. These results were also replicated in sensitivity analyses that incorporated echocardiogram measurements into clinical outcome definitions, further validating our primary definition (eResults, eTable 5). These strong associations were also consistent in sensitivity analyses restricted to individuals genetically similar to a European reference population, and diminished in analyses restricted to individuals genetically similar to an African reference population (eTables 6-9). These findings demonstrate that PGSs for HCM and DCM are significantly and bidirectionally associated with both HCM and DCM in the PMBB population, and specifically in individuals genetically similar to the European populations in which the genomic data used to develop the PGS weights were originally derived.

**Figure 3:**
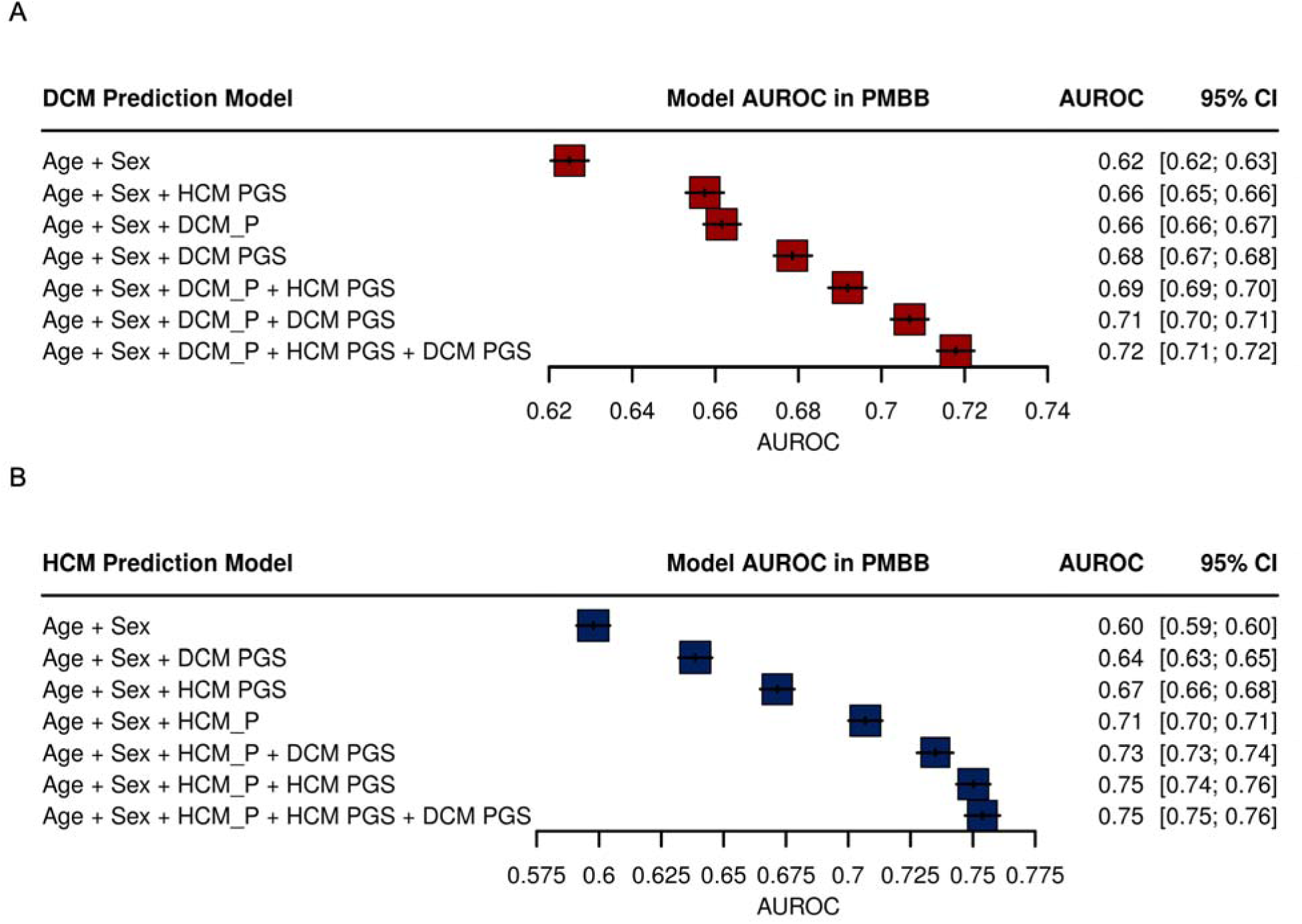
Contributions of monogenic and polygenic risk terms to A) DCM and B) HCM model discrimination. HCM_P and DCM_P indicate the presence of a pathogenic variant in an HCM and DCM gene respectively.

Because abnormal cardiac morphologies are hallmark features of HCM and DCM, we next tested for subclinical associations between the PGSs and clinically obtained echocardiographic measurements. A one standard deviation increase in HCM PGS was associated with a 0.18 mm increase in IVS (95% CI 0.14-0.22; p=9.3x10^-19^); the DCM PGS did not have a significant association with IVS (estimate -1.3 x10^-3^ mm; 95% CI -0.036- -0.034; p=0.94). A one standard deviation increase in HCM PGS was associated with an increase in ejection fraction of 1.0% (95% CI 0.87-1.3; p = 6.8x10^-28^). A one standard deviation increase in DCM PGS was associated with a decrease in ejection fraction of 1.9% (95% CI -2.2 to -1.7; p = 1.5x10^-62^). A one standard deviation increase in HCM PGS was associated with a 0.79 mm decrease in LVIDd (95% CI -0.92 to -0.67; p = 2.3x10^-36^), and a one standard deviation increase in DCM PGS was associated with a 1.0 mm increase in LVIDd (95% CI 0.93-1.1; p = 3.2x10^-78^). These results demonstrate that PGSs are associated with small but significant differences in echocardiographic measures relevant to HCM and DCM.

### Polygenic Modification of Monogenic Risk

We next sought to evaluate whether polygenic background bidirectionally modifies the penetrance of monogenic variants in HCM and DCM. We modeled each disease outcome (HCM or DCM) as a function of monogenic pathogenic variant status and polygenic risk, including an interaction term, and adjusting for covariates. We observed significant independent associations of monogenic variation and either an HCM or DCM PGS). Interaction terms between polygenic and monogenic risk were not significant (Figure 2, eTables 6-9).

To assess how each of these monogenic and polygenic variables individually or collectively contributed to disease risk we assessed the discrimination and calibration of predictive models that additively included each term. Calibration of the HCM model, characterized by Brier score, was only improved by consideration of HCM pathogenic variant status, and the DCM model was not markedly improved by inclusion of monogenic and polygenic model terms (eFigure 4). However, discrimination, measured with AUROC, of models predicting prevalent HCM and DCM were each significantly (>95% probability of a difference) improved when considering terms for both monogenic and polygenic risk, compared to a baseline model including age and sex (Figure 3). The discrimination of an HCM model was improved more by the addition of an HCM PGS than a DCM PGS, and the opposite was true in a DCM model; nevertheless, both HCM and DCM PGSs conferred significant improvement for models of either outcome alone or in addition to a monogenic risk term. The performance improvement for an age- and sex-only DCM model was improved more by a DCM PGS (mean of posterior distribution of AUROC difference 0.054; 95% credible interval 0.047-0.060) than by a monogenic DCM risk term (mean of posterior distribution of AUROC difference 0.037; 95% credible interval 0.030-0.043).

**Figure 4:**
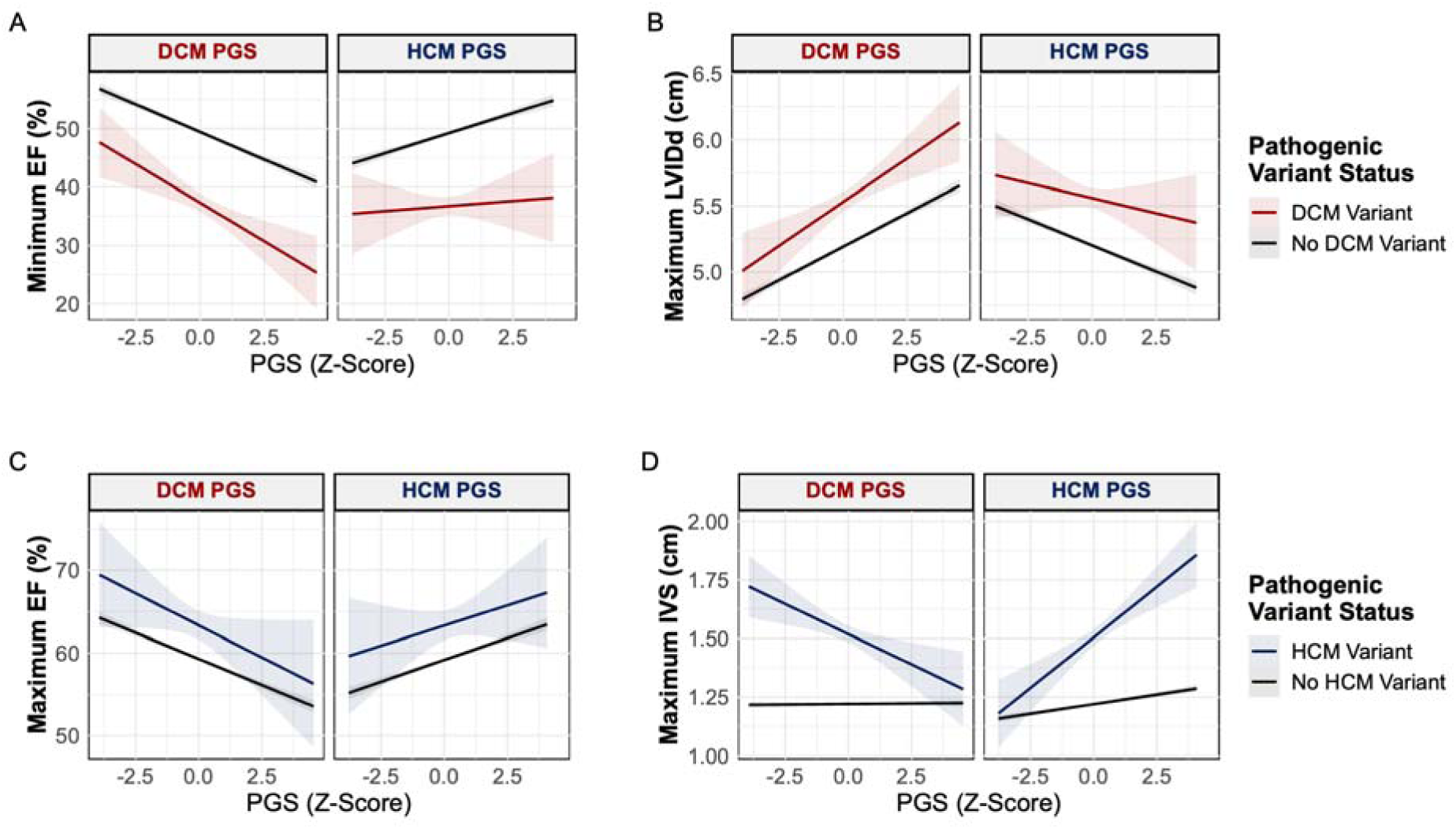
Effects of polygenic and monogenic risk on echocardiogram measurements. Linear regression models including polygenic risk, monogenic risk, a polygenic:monogenic interaction term, sex, and age at time of echocardiogram were used for marginal prediction of echocardiogram measurements. **A)** Predicted Minimum EF and **B)** Predicted Maximal LVIDd stratified by DCM pathogenic variant status using DCM (left panel) and HCM (right panel) PGSs. **C)** Predicted Maximum EF and **D)** Predicted Maximal IVS stratified by HCM pathogenic variant status using DCM (left panel) and HCM (right panel) PGSs.

The combined effects of polygenic and monogenic background on echocardiogram measurements was also assessed. In most cases, the PGS contributed additively to the baseline effect associated with carrying a pathogenic variant, with HCM and DCM PGSs exhibiting opposite effects (Figure 4, eTables 10-13). Notably, for IVS we observed significant (p < 0.01) interaction terms between HCM monogenic risk and either the HCM or DCM polygenic scores, suggesting that the influence of monogenic variants on septal thickness may be uniquely modified by polygenic background (eTable 10).

## Discussion

In this study, we leveraged data from a large, diverse biobank to characterize both monogenic and polygenic contributions to HCM and DCM. First, we affirmed that pathogenic variants in established HCM and DCM genes are strongly associated with their respective condition. Next, we identified bidirectional associations between HCM and DCM PGSs and their corresponding disease diagnosis as well as characteristic echocardiographic features. Finally, we found that polygenic background bidirectionally modifies the penetrance of monogenic variants. Predictive models benefited from the inclusion of both polygenic risk and monogenic risk terms, indicating the independent importance of both rare and common variant background to disease liability. These findings add to the growing body of evidence that HCM and DCM exist on opposite ends of an overlapping genotypic and phenotypic spectrum, demonstrate the bidirectional impact of polygenic background on pathogenic variant penetrance, and carry implications for clinical genetic testing.

We establish that beyond individual polymorphisms, common variant background in aggregate impacts inherited cardiomyopathy along a spectrum. Cardiac sarcomere function is known to play a critical role in both HCM and DCM through opposing mechanisms that result in increased or decreased contractility, respectively.^3^ Common variants at a limited number of loci have previously been implicated as bidirectional contributors to HCM and DCM, and we support and extend these findings by demonstrating that PGS for these conditions have opposing phenotypic manifestations.^18,19^ The bi-directional associations of the HCM and DCM PGSs with clinical diagnoses and subclinical echocardiographic measures suggests these inherited cardiomyopathies exist as pathologic extremes of a genetic and phenotypic continuum.

Our finding that the penetrance of pathogenic variants for HCM and DCM is bidirectionally modified by polygenic background adds additional nuance to our understanding of inherited cardiomyopathy risk. Viewing HCM and DCM through the lens of the ‘liability threshold’ model of disease (which posits that disease manifests when the cumulative contributions of risk factors reach a critical threshold), previous studies have separately established in HCM and DCM that polygenic risk influences the likelihood that individuals with and without monogenic risk cross this critical threshold.^30^ Our work demonstrates that these thresholds are not independent; the same polygenic background either increase or decrease cardiomyopathy susceptibility, depending on carrier status for pathogenic variants that predispose to HCM or DCM. Notably, the effects of HCM and DCM PGSs in modifying monogenic risk were not perfect mirror images, and these traits are only moderately correlated at the genetic level. Defining the shared and unique loci contributing to these inherited cardiomyopathies is an area of active investigation that may help define new biological mechanisms contributing to the phenotypic spectrum of disease.^14,15,18^

These results have implications for how clinical genetic testing for inherited cardiomyopathies is considered in the future. Current practice guidelines for cardiomyopathy leverage panel genetic testing to stratify risk based on the presence or absence of typically rare pathogenic variants, broadly recommending serial clinical screening in unaffected variant carriers, and cascade genetic testing of family members when a pathogenic variant is identified.^31^ Yet – as our study and others have identified – these rare pathogenic variants have incomplete penetrance, such that many individuals subjected to this screening will not develop disease. In the future, additional consideration of polygenic background may inform more precise recommendations. For example, those with favorable polygenic background may undergo less frequent clinical screening, and polygenic background may contribute to assessment of disease expressivity/severity. Clinical representation of polygenic risk could take the form of an isolated measurement, or alternatively part of an integrated risk score akin to those developed for other cardiovascular diseases such as coronary artery disease.^32^ Given the rapid growth of targeted molecular therapies for inherited cardiomyopathies, PGSs may have a role in enriching clinical trial populations for those at higher risk who might derive the greatest benefit from these novel prevention and treatment strategies.^33^ While our results suggest that cardiomyopathy PGSs improve discrimination of inherited cardiomyopathy prediction models at the population level, they are not ready for use in clinical decision-making, and rigorous prospective clinical validation across broad population subgroups will be an essential step to justify any individual-level implementation.^34^

### Limitations

Our study has several limitations. First, HCM and DCM are modestly genetically correlated at the summary and individual levels, indicating that some bidirectional associations of HCM and DCM PGSs are attributable to shared genetic signals with opposing directions of effect. Further investigation to detangle shared and distinct components of HCM and DCM polygenic background is warranted. Second, carriers of pathogenic variants in *MYH7, TNNC1, PLN* and *TNNT2* were considered in both the HCM and DCM monogenic risk groups because these genes have strong evidence of pathogenicity for both HCM and DCM. Including these carriers in both risk groups should bias associations toward the null, making the overall trends of opposing direction of risk modification all the more striking. Third, an additional consequence of grouping carriers of pathogenic variants in HCM and DCM genes is that we are unable to assess if modification through polygenic background differs by specific gene. Fourth, we considered polygenic risk as a continuous variable consistent with the well-established liability threshold model, although it is possible that genetic susceptibility is conferred in more discrete units. The additive approach of PGSs also may underestimate the true impact of polygenic risk by failing to account for complex variant relationships such as epistasis.^35^ Finally, the previously published genomic studies used to generate the polygenic scores were carried out in individuals genetically similar to European reference populations. While the relationship between polygenic background and monogenic variant status should be broadly relevant, the existing PGSs may not be generalizable across populations, as seen in sensitivity analyses in individuals genetically similar to an African reference population. We anticipate that future large GWAS of inherited cardiomyopathies among broader populations will improve the transportability of PGSs and facilitate broader generalization of our results.

## Conclusions

Our study demonstrates that polygenic scores for HCM and DCM associate with clinical and echocardiographic measures relevant to both diseases, and polygenic background bidirectionally modifies the penetrance of pathogenic variation in genes definitively associated with inherited cardiomyopathies. Polygenic background for HCM and DCM exists on opposite ends of an overlapping spectrum.

## Data Availability

The DCM PGS will be deposited on the Polygenic Score Catalog upon manuscript publication.

## Acknowledgements

We acknowledge the Penn Medicine BioBank (PMBB) for providing data and thank the patient-participants of Penn Medicine who consented to participate in this research program. We also thank the Penn Medicine BioBank team and Regeneron Genetics Center for providing genetic variant data for analysis. The PMBB is approved under IRB protocol# 813913 and supported by Perelman School of Medicine at University of Pennsylvania, a gift from the Smilow family, and the National Center for Advancing Translational Sciences of the National Institutes of Health under CTSA award number UL1TR001878.

