## Supplemental Material for "Polygenic Background and Penetrance of Pathogenic Variants in Hypertrophic and Dilated Cardiomyopathies"

Supplemental Content

### **eMethods**

Validation of Clinical Outcome Definitions with Echocardiogram Data

Associations between clinical outcome definitions and echocardiogram measurements were assessed with Wilcoxon rank-sum tests. We additionally assessed the proportion of cases meeting diagnosis code-based definitions also meeting commonly used echocardiographic thresholds (LVEF <50% for DCM, maximum IVS >1.5 cm for HCM).^1,2^

Population Group Assignment

Population group assignment was determined using *pgsc_calc* genetic similarity analysis within its ANCESTRY_ANALYSIS module and tool of its ‘pgscatalog_utils’ package. To identify the population that an individual is most genetically similar to, a RandomForest classifier is trained to classify samples based on their PC-loadings to the most genetically similar population in the provided 1KG + HGDP reference panel: African (AFR), admixed American (AMR), East Asian (EAS), European (EUR), Middle Eastern (MID), and Central/South Asian (CSA).

Polygenic Score Generation

At the time of analysis, the published DCM PGSs with the highest reported performance (Jurgens et al.) did not include the full DCM GWAS cohort. We therefore used PRS-CSx to create a new score that uses the full GWAS summary statistic data. Effective sample size was calculated as 4/((1/ncases) + (1/ncontrols)).

### **eResults**

Selection of Study Population

Starting with 49,434 unrelated PMBB participants with genotypes and available diagnosis codes, for analyses that used cardiomyopathy clinical outcomes the study population was further filtered based on the diagnosis codes listed in eTable 1.

828 individuals had an HCM ‘phenocopy’ code and 93 had both HCM and DCM inclusion codes. 8,123 participants had a heart failure diagnosis code but did not meet the HCM or DCM case definition criteria (eTable 1). This filtering resulted in a study population of 419 HCM cases, 895 DCM cases, and 39,421 non-cases.

Validation of Clinical Outcome Definitions

Of the 365 individuals meeting our HCM clinical outcome definition with available IVS measurements, 225 had a maximum IVS of >1.5 cm. This number increased to 293 with an adjusted cutoff of >1.3 cm used in the presence of a family history. Of the 816 individuals meeting DCM clinical outcome definitions with available EF measurements, 714 had a minimum recorded EF of <= 50%. This demonstrated concordance between imaging and code-based diagnostic definitions.

As a sensitivity analysis to explore the possibility of phenotypic misclassification attenuating observed associations, we tested associations between polygenic scores and cardiomyopathy outcomes that used more stringent HCM and DCM definitions, requiring participants to meet clinical outcome definitions as well as the aforementioned measurement cutoffs. All estimates of associations between PGSs and these more stringent outcomes fell within the 95% confidence intervals of the estimates for the primary outcome definitions, further validating this definition (original results found in main manuscript, sensitivity analysis results in eTable 5).

### **eTables**

#### **eTable 1: Diagnosis-Code Based Definitions.**

| **Disease** | **ICD Codes** | **CPT Codes** | **Exclusions** |
| --- | --- | --- | --- |
| All Cause Heart Failure (HF) | I11.0, I13.0, I13.2, I25.5, I42.0, I42.1, I42.2, I42.6, I42.7, I50, O90.3  425.1, 425.11, 425.18, 425.2, 425.9, 428, 674.5 |  |  |
| Atrial Fibrillation (AF) | 427.3, 427.31, I48.91, I48.0, I48.2, I48, I48.1 |  |  |
| Hypertension (HTN) | 401.0, 401.1, 401.9, I10 |  |  |
| Aortic Stenosis (AS) | 424.1(2+) OR I35.0 | 33361-33369, 35.05, 35.06, 25.21, 35.22, O2RF, 33405-33410, 33411-33413 | ICD9: 746,747  ICD10: Q20-26 |
| Coronary Artery Disease (CAD) | I21, I22, I23, I24, I25.2  410, 411, 412 | 33510, 33511, 33512, 33513, 33533, 33534, 33535, 33536, 92920, 92921, 92924, 92925, 92928, 92929, 92933, 92934, 92937, 92938, 92941, 92943, 92944, 92973, 92975 |  |
| Dilated Cardiomyopathy | I42.0 |  | CAD prior to or on date of DCM code, co-existence of HCM code |
| Hypertrophic Cardiomyopathy (HCM) | I42.1, I42.2, 425.18, 425.11, 425.1 |  | Any HCM phenocopy code, co-existence of DCM code |
| HCM Phenocopy |  |  |  |
| Fabry Disease | E75.21 |  |  |
| Amyloidosis | 277.3, 277.30, 277.39, E85.0, E85.1, E85.2, E85.3, E85.4, E85.8, E85.81, E85.82, E85.89, E85.9 |  |  |
| Fabry Disease | E75.21 |  |  |
| Hemochromatosis | 275.01, 275.03, E83.110, E83.118, E83.119 |  |  |
| Glycogen Storage Diseases | 271.0, 271.00, 271.01, 271.02, 271.03, 271.04, 271.09, E74.00, E74.01, E74.02, E74.03, E74.04, E74.09 |  |  |
| LAMP2/Danon Disease | E75.4 |  |  |
| Mitochondrial Disorders | 277.87, E88.40, E88.41, E88.42, E88.49 |  |  |
| Noonan Syndrome | 759.89, Q87.1 |  |  |
| Friedreich's Ataxia | 334.0, G11.1 |  |  |
| Fatty Acid Oxidation Disorders | 277.85, 277.81, 277.82, E71.310, E71.311, E71.312, E71.318, E71.42 |  |  |
| Other Metabolic Disorders | 270.0, 270.1, 270.2, 270.3, 270.4, 270.5, 270.6, 270.7, 270.8, 270.9, 271.1, 271.11, 277.5, 277.51, 277.52, 277.53, 277.59, 277.86, E72.5, E77.0, E77.1, E77.8, E77.9 |  |  |

AS diagnosis requires code at age >= 55. In HCM specifically, case status requires two diagnosis codes recorded on different dates. DCM and HCM non-cases are defined by the absence of any HCM, DCM, HF, or ‘HCM Phenocopy’ inclusion code.

#### **eTable 2: Baseline Characteristics of Study Participants.**

|  | **HCM**  (n = 419) | **DCM**  (n = 895) | **Non-case**  (n = 39,421) |
| --- | --- | --- | --- |
| Age, mean (SD), y | 58.5 (14.9) | 56.7 (13.9) | 52.7 (16.5) |
| Sex, No. (%) |  |  |  |
| Male | 221 (52.7) | 610 (68.2) | 18,598 (47.2) |
| Female | 198 (47.3) | 285 (31.8) | 20,823 (52.8) |
| Population, No. (%) |  |  |  |
| European | 308 (73.5) | 605 (67.6) | 28,846 (73.2) |
| African | 91 (21.7) | 238 (26.6) | 7,609 (19.3) |
| Other | 20 (4.8) | 52 (5.8) | 2,966 (7.5) |
| BMI, mean (SD)  n = 39,157 | 30.3 (6.3) | 30.3 (6.8) | 29.1 (6.6) |
| Hypertension Diagnosis, No. (%) | 350 (83.5) | 764 (85.4) | 21,487 (54.5) |
| Coronary Artery Disease, No. (%) | 100 (23.9) | 120 (13.4) | 3,496 (8.9) |
| Atrial Fibrillation, No. (%) | 237 (56.6) | 572 (63.9) | 6,372 (16.2) |
| Calcific Aortic Stenosis, No. (%) | 57 (13.6) | 80 (8.9) | 1,484 (3.8) |

Abbreviations: BMI, body mass index (calculated as weight in kilograms divided by height in meters squared); HCM, hypertrophic cardiomyopathy; DCM, dilated cardiomyopathy. Population categorization was determined based on genetic similarity to an external reference population. “Other” includes Admixed American, Central/South Asian, East Asian, and Middle Eastern reference populations.

#### **eTable 3: Baseline Characteristics, Stratified by Echocardiogram Status.**

|  | **With Echo**  (n = 21,674) | **Without Echo**  (n = 27,760) |
| --- | --- | --- |
| Age, mean (SD), y | 59.1 (14.9) | 51.0 (16.8) |
| Sex, No. (%) |  |  |
| Male | 12,236 (56.5) | 12,650 (45.6) |
| Female | 9,438 (43.5) | 15,110 (54.4) |
| Population, No. (%) |  |  |
| European | 15,729 (72.6) | 20,097 (72.4) |
| African | 4,807 (22.2) | 5,370 (19.3) |
| Other | 1,138 (5.3) | 2,293 (8.3) |
| BMI, mean (SD)  n = 47,688 | 29.5 (6.6) | 29.1 (6.7) |
| Hypertension Diagnosis, No. (%) | 17,038 (78.6) | 13,199 (47.5) |
| Coronary Artery Disease, No. (%) | 5,605 (25.9) | 1,724 (6.2) |
| Atrial Fibrillation, No. (%) | 9,410 (43.4) | 2 2,779 (10.0) |
| Calcific Aortic Stenosis, No. (%) | 2,580 (11.9) | 550 (2.0) |

Abbreviations: BMI, body mass index (calculated as weight in kilograms divided by height in meters squared). Population categorization was determined based on genetic similarity to an external reference population. “Other” includes Admixed American, Central/South Asian, East Asian, and Middle Eastern reference populations.

#### **eTable 4: Pathogenic Variation in Cardiomyopathy Cases**

|  | **HCM Case** (n = 419) | **DCM Case** (n = 895) | **Non-Case** (n = 39,421) |
| --- | --- | --- | --- |
| HCM Pathogenic Variant, No. (%) | 84 (20) | 6 (0.7) | 169 (0.4) |
| DCM Pathogenic Variant, No. (%) | 45 (11) | 94 (11) | 349 (0.9) |

Counts of HCM and DCM pathogenic variant carriers among study participants stratified by their HCM, DCM, and non-case status.

#### **eTable 5: Associations of PGSs with HCM and DCM, defined using EHR codes and echocardiogram measurements**

|  | **HCM** | **DCM** |
| --- | --- | --- |
| HCM PGS | OR 1.8; 95% CI 1.6-2.1; p = 3.7x10^-15^ | OR 0.64; 95% CI 0.59-0.70; p = 5.3x10^-23^ |
| DCM PGS | OR 0.66; 95% CI 0.57-0.75; p = 5.9x10^-10^ | OR 1.6; 95% CI 1.5-1.8; p = 4.3x10^-35^ |

DCM was defined as meeting the primary diagnosis code-based definition in addition to minimum measured LVEF <= 50%. HCM was defined as meeting the primary diagnosis code-based definition in addition to measured IVS of >= 1.5 cm. Models include sex and age at enrollment as covariates.

#### **eTable 6: Population-Stratified Models of HCM Including HCM PGS and HCM Pathogenic Variant Status**

|  | **Population** | | |
| --- | --- | --- | --- |
|  | **ALL** | **EUR** | **AFR** |
| Intercept | 0.00 (0.00-0.00), p=2.91e-207 | 0.00 (0.00-0.00), p=1.43e-136 | 0.00 (0.00-0.00), p=9.33e-53 |
| HCM PGS | 1.75 (1.55-1.98), p=1.94e-19 | 1.91 (1.65-2.22), p=6.35e-18 | 1.29 (1.01-1.64), p=0.038 |
| HCM Pathogenic | 63.77 (45.91-87.58), p=6.55e-141 | 76.92 (52.91-110.45), p=5.71e-119 | 23.30 (8.38-55.33), p=2.53e-11 |
| Age | 1.03 (1.02-1.03), p=2.03e-14 | 1.02 (1.01-1.03), p=5.52e-08 | 1.04 (1.03-1.06), p=1.20e-07 |
| Sex | 1.07 (0.88-1.32), p=0.495 | 1.06 (0.84-1.36), p=0.611 | 1.32 (0.86-2.01), p=0.204 |
| HCM PGS: HCM Pathogenic | 1.02 (0.74-1.43), p=0.896 | 1.05 (0.72-1.56), p=0.811 | 0.79 (0.34-1.89), p=0.588 |

Results of population-stratified logistic regression models of HCM, with model terms including a HCM PGS, HCM pathogenic variant status (‘HCM Pathogenic’), age at PMBB enrollment, sex, and an interaction term between HCM PGS and HCM pathogenic variant status (‘HCM PGS: HCM Pathogenic’). Results are displayed with odds ratios, 95% confidence intervals, and p-values, and are shown for all-comers (‘ALL’), individuals genetically similar to a European reference population (‘EUR’), and individuals genetically similar to an African reference population (‘AFR’).

#### **eTable 7: Population-Stratified Models of DCM Including DCM PGS and DCM Pathogenic Variant Status**

|  | **ALL** | **EUR** | **AFR** |
| --- | --- | --- | --- |
| Intercept | 0.01 (0.00-0.01), p=0.00e+00 | 0.00 (0.00-0.01), p=2.75e-207 | 0.01 (0.01-0.02), p=7.26e-79 |
| DCM PGS | 1.59 (1.48-1.71), p=7.35e-36 | 1.73 (1.58-1.89), p=9.44e-32 | 1.38 (1.21-1.57), p=1.43e-06 |
| DCM Pathogenic | 13.01 (9.94-16.82), p=1.20e-81 | 15.99 (11.94-21.13), p=5.41e-81 | 4.52 (1.21-11.71), p=0.007 |
| Age | 1.01 (1.01-1.02), p=1.25e-07 | 1.01 (1.01-1.02), p=1.55e-06 | 1.01 (1.00-1.02), p=0.014 |
| Sex | 2.21 (1.91-2.55), p=2.61e-26 | 2.24 (1.87-2.70), p=5.43e-18 | 2.69 (2.06-3.54), p=8.07e-13 |
| DCM PGS: DCM Pathogenic | 1.00 (0.78-1.28), p=0.978 | 0.84 (0.65-1.11), p=0.212 | 2.51 (1.10-7.22), p=0.049 |

Results of population-stratified logistic regression models of DCM, with model terms including a DCM PGS, DCM pathogenic variant status (‘DCM Pathogenic’), age at PMBB enrollment, sex, and an interaction term between DCM PGS and DCM pathogenic variant status (‘DCM PGS: DCM Pathogenic’). Results are displayed with odds ratios, 95% confidence intervals, and p-values, and are shown for all-comers (‘ALL’), individuals genetically similar to a European reference population (‘EUR’), and individuals genetically similar to an African reference population (‘AFR’).

#### **eTable 8: Population-Stratified Models of DCM Including HCM PGS and DCM Pathogenic Variant Status**

|  | **ALL** | **EUR** | **AFR** |
| --- | --- | --- | --- |
| Intercept | 0.01 (0.01-0.01), p=0.00e+00 | 0.00 (0.00-0.01), p=1.71e-206 | 0.01 (0.01-0.02), p=1.52e-77 |
| HCM PGS | 0.67 (0.62-0.73), p=5.09e-22 | 0.64 (0.58-0.71), p=1.82e-17 | 0.68 (0.58-0.79), p=4.24e-07 |
| DCM Pathogenic | 13.58 (10.59-17.27), p=3.29e-97 | 16.00 (12.15-20.88), p=7.53e-90 | 7.77 (3.45-15.72), p=7.92e-08 |
| Age | 1.01 (1.01-1.02), p=1.49e-07 | 1.01 (1.01-1.02), p=1.05e-06 | 1.01 (1.00-1.02), p=0.013 |
| Sex | 2.18 (1.89-2.53), p=1.01e-25 | 2.24 (1.87-2.69), p=7.00e-18 | 2.66 (2.04-3.50), p=1.07e-12 |
| HCM PGS: DCM Pathogenic | 1.23 (0.91-1.66), p=0.174 | 1.23 (0.87-1.73), p=0.233 | 1.46 (0.63-3.31), p=0.360 |

Results of population-stratified logistic regression models of DCM, with model terms including a HCM PGS, DCM pathogenic variant status (‘DCM Pathogenic’), age at PMBB enrollment, sex, and an interaction term between HCM PGS and DCM pathogenic variant status (‘HCM PGS: DCM Pathogenic’). Results are displayed with odds ratios, 95% confidence intervals, and p-values, and are shown for all-comers (‘ALL’), individuals genetically similar to a European reference population (‘EUR’), and individuals genetically similar to an African reference population (‘AFR’).

#### **eTable 9: Population-Stratified Models of HCM Including DCM PGS and HCM Pathogenic Variant Status**

|  | **ALL** | **EUR** | **AFR** |
| --- | --- | --- | --- |
| Intercept | 0.00 (0.00-0.00), p=1.76e-206 | 0.00 (0.00-0.00), p=3.09e-139 | 0.00 (0.00-0.00), p=4.24e-52 |
| DCM PGS | 0.68 (0.61-0.76), p=4.07e-12 | 0.56 (0.49-0.64), p=3.99e-18 | 1.03 (0.84-1.28), p=0.756 |
| HCM Pathogenic | 68.23 (50.37-91.86), p=1.59e-167 | 91.77 (64.94-129.05), p=2.80e-147 | 21.73 (8.14-50.50), p=1.75e-11 |
| Age | 1.03 (1.02-1.03), p=5.39e-14 | 1.02 (1.01-1.03), p=1.06e-07 | 1.04 (1.03-1.06), p=9.71e-08 |
| Sex | 1.07 (0.88-1.32), p=0.486 | 1.08 (0.85-1.38), p=0.509 | 1.31 (0.86-2.01), p=0.208 |
| DCM PGS: HCM Pathogenic | 1.35 (1.01-1.80), p=0.041 | 1.58 (1.12-2.22), p=0.008 | 0.76 (0.37-1.53), p=0.436 |

Results of population-stratified logistic regression models of HCM, with model terms including a DCM PGS, HCM pathogenic variant status (‘HCM Pathogenic’), age at PMBB enrollment, sex, and an interaction term between DCM PGS and HCM pathogenic variant status (‘DCM PGS: HCM Pathogenic’). Results are displayed with odds ratios, 95% confidence intervals, and p-values, and are shown for all-comers (‘ALL’), individuals genetically similar to a European reference population (‘EUR’), and individuals genetically similar to an African reference population (‘AFR’).

#### **eTable 10: Septal Thickness as a Function of Pathogenic Variant Status and either HCM or DCM PGS**

|  | **HCM PGS** | **DCM PGS** |
| --- | --- | --- |
| Intercept | 0.85 (0.84-0.87), p=0.00e+00 | 0.86 (0.84-0.87), p=0.00e+00 |
| PGS | 0.02 (0.01-0.02), p=4.97e-16 | 0.00 (-0.00-0.00), p=0.607 |
| HCM Pathogenic | 0.29 (0.25-0.32), p=3.75e-51 | 0.30 (0.26-0.33), p=2.58e-57 |
| Age | 0.00 (0.00-0.00), p=3.48e-240 | 0.00 (0.00-0.00), p=9.18e-239 |
| Sex | 0.13 (0.12-0.13), p=2.39e-262 | 0.13 (0.12-0.13), p=8.06e-261 |
| PGS: HCM Pathogenic | 0.07 (0.03-0.11), p=1.09e-04 | -0.05 (-0.09--0.02), p=0.002 |

Results of linear regression models of interventricular septal thickness (measured in centimeters), with model terms including either and HCM or DCM PGS, HCM pathogenic variant status (‘HCM Pathogenic’), age at the time of echocardiogram, sex, and an interaction term between the PGS and HCM pathogenic variant status (‘PGS: HCM Pathogenic’). Results are displayed with beta coefficients, 95% confidence intervals, and p-values.

#### **eTable 11: LVEF as a function of HCM pathogenic variants status and either HCM or DCM PGS**

|  | **HCM PGS** | **DCM PGS** |
| --- | --- | --- |
| Intercept | 62.96 (62.26-63.66), p=0.00e+00 | 63.19 (62.49-63.89), p=0.00e+00 |
| PGS | 1.06 (0.86-1.25), p=3.64e-27 | -1.26 (-1.43--1.10), p=3.90e-49 |
| HCM Pathogenic | 4.16 (2.37-5.96), p=5.43e-06 | 4.06 (2.30-5.81), p=5.80e-06 |
| Age | 0.01 (-0.01-0.02), p=0.359 | 0.00 (-0.01-0.01), p=0.533 |
| Sex | -4.11 (-4.45--3.77), p=3.07e-122 | -4.11 (-4.45--3.77), p=1.16e-122 |
| PGS: HCM Pathogenic | -0.08 (-1.79-1.63), p=0.928 | -0.29 (-1.91-1.33), p=0.727 |

Results of linear regression models of left ventricular ejection fraction with model terms including either and HCM or DCM PGS, HCM pathogenic variant status (‘HCM Pathogenic’), age at the time of echocardiogram, sex, and an interaction term between the PGS and HCM pathogenic variant status (‘PGS: HCM Pathogenic’). Results are displayed with beta coefficients, 95% confidence intervals, and p-values.

#### **eTable 12: LVIDD as a function of DCM pathogenic variants status and either HCM or DCM PGS**

|  | **HCM PGS** | **DCM PGS** |
| --- | --- | --- |
| Intercept | 4.83 (4.78-4.87), p=0.00e+00 | 4.84 (4.80-4.89), p=0.00e+00 |
| PGS | 0.10 (0.09-0.11), p=1.60e-73 | -0.08 (-0.09--0.07), p=2.00e-35 |
| DCM Pathogenic | 0.34 (0.26-0.42), p=1.23e-17 | 0.36 (0.28-0.43), p=4.45e-20 |
| Age | -0.00 (-0.00--0.00), p=4.20e-26 | -0.00 (-0.00--0.00), p=2.40e-27 |
| Sex | 0.60 (0.58-0.62), p=0.00e+00 | 0.60 (0.58-0.62), p=0.00e+00 |
| PGS: DCM Pathogenic | 0.03 (-0.04-0.10), p=0.370 | 0.03 (-0.05-0.12), p=0.454 |

Results of linear regression models of left ventricular internal end diastolic diameter (measured in centimeters), with model terms including either and HCM or DCM PGS, DCM pathogenic variant status (‘DCM Pathogenic’), age at the time of echocardiogram, sex, and an interaction term between the PGS and DCM pathogenic variant status (‘PGS: DCM Pathogenic’). Results are displayed with beta coefficients, 95% confidence intervals, and p-values.

#### **eTable 13: LVEF as a function of DCM pathogenic variants status and either HCM or DCM PGS**

|  | **HCM PGS** | **DCM PGS** |
| --- | --- | --- |
| Intercept | 55.38 (54.44-56.33), p=0.00e+00 | 55.69 (54.75-56.63), p=0.00e+00 |
| PGS | 1.37 (1.11-1.64), p=4.76e-25 | -1.88 (-2.11--1.65), p=5.07e-58 |
| DCM Pathogenic | -12.49 (-14.07--10.91), p=1.01e-53 | -12.12 (-13.72--10.52), p=1.39e-49 |
| Age | 0.00 (-0.01-0.02), p=0.883 | -0.00 (-0.02-0.01), p=0.880 |
| Sex | -6.21 (-6.67--5.75), p=6.95e-151 | -6.20 (-6.66--5.74), p=1.82e-151 |
| PGS: DCM Pathogenic | -1.03 (-2.86-0.80), p=0.269 | -0.75 (-2.16-0.67), p=0.300 |

Results of linear regression models of left ventricular ejection fraction with model terms including either and HCM or DCM PGS, DCM pathogenic variant status (‘DCM Pathogenic’), age at the time of echocardiogram, sex, and an interaction term between the PGS and DCM pathogenic variant status (‘PGS: DCM Pathogenic’). Results are displayed with beta coefficients, 95% confidence intervals, and p-values.

### **eFigures**

#### **eFigure 1: Validation of Cardiomyopathy Diagnosis Definitions with Echocardiogram Measurements**

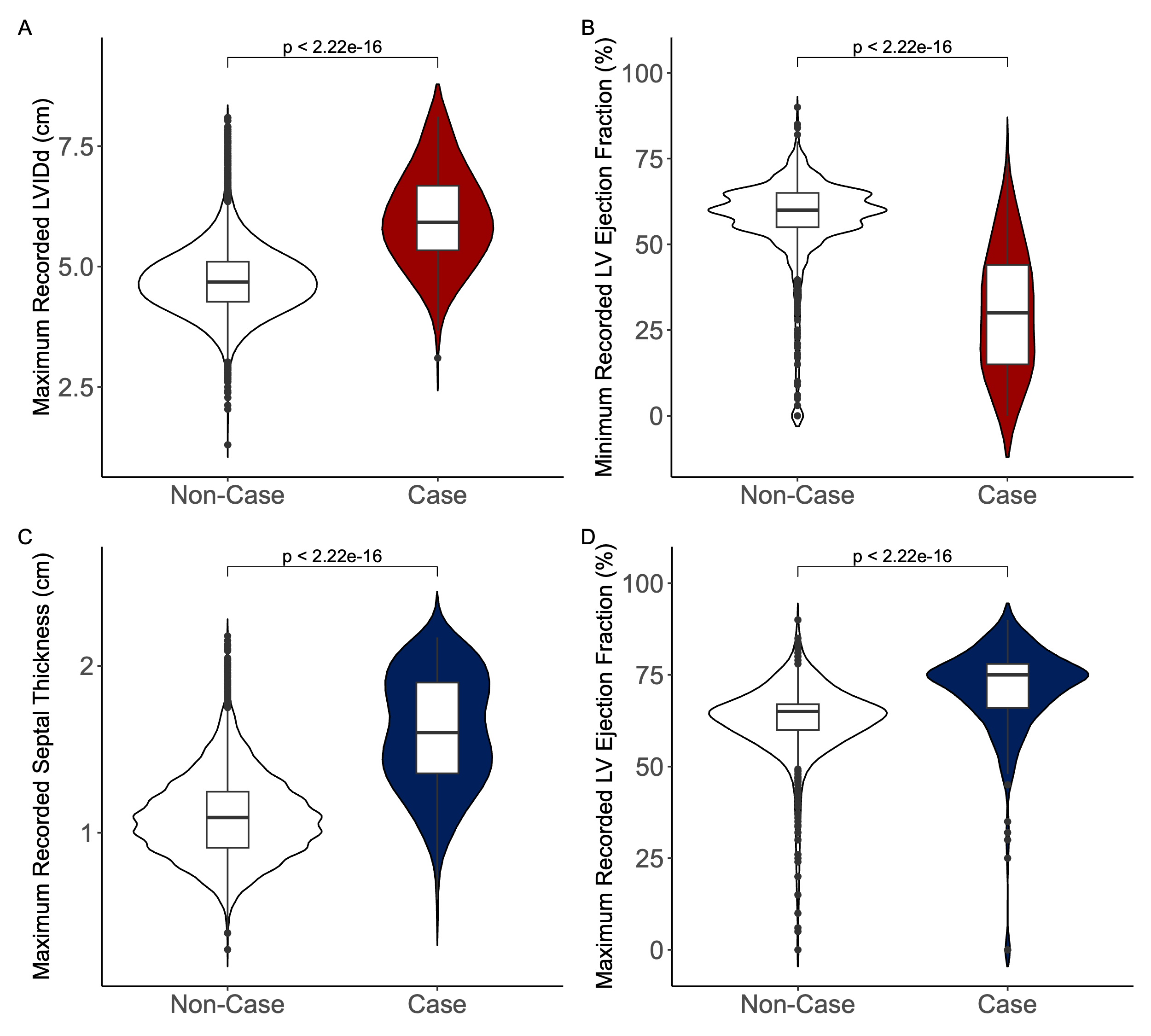

**A)** Maximum recorded LVIDd in DCM cases vs non-cases; **B)** Minimum recorded LVEF in DCM cases vs non-cases **C)** Maximum recorded IVS in HCM cases vs non-cases; **D)** Maximum recorded LVEF in HCM cases vs non-cases.

#### **eFigure 2: Cardiomyopathy Cases by Gene**

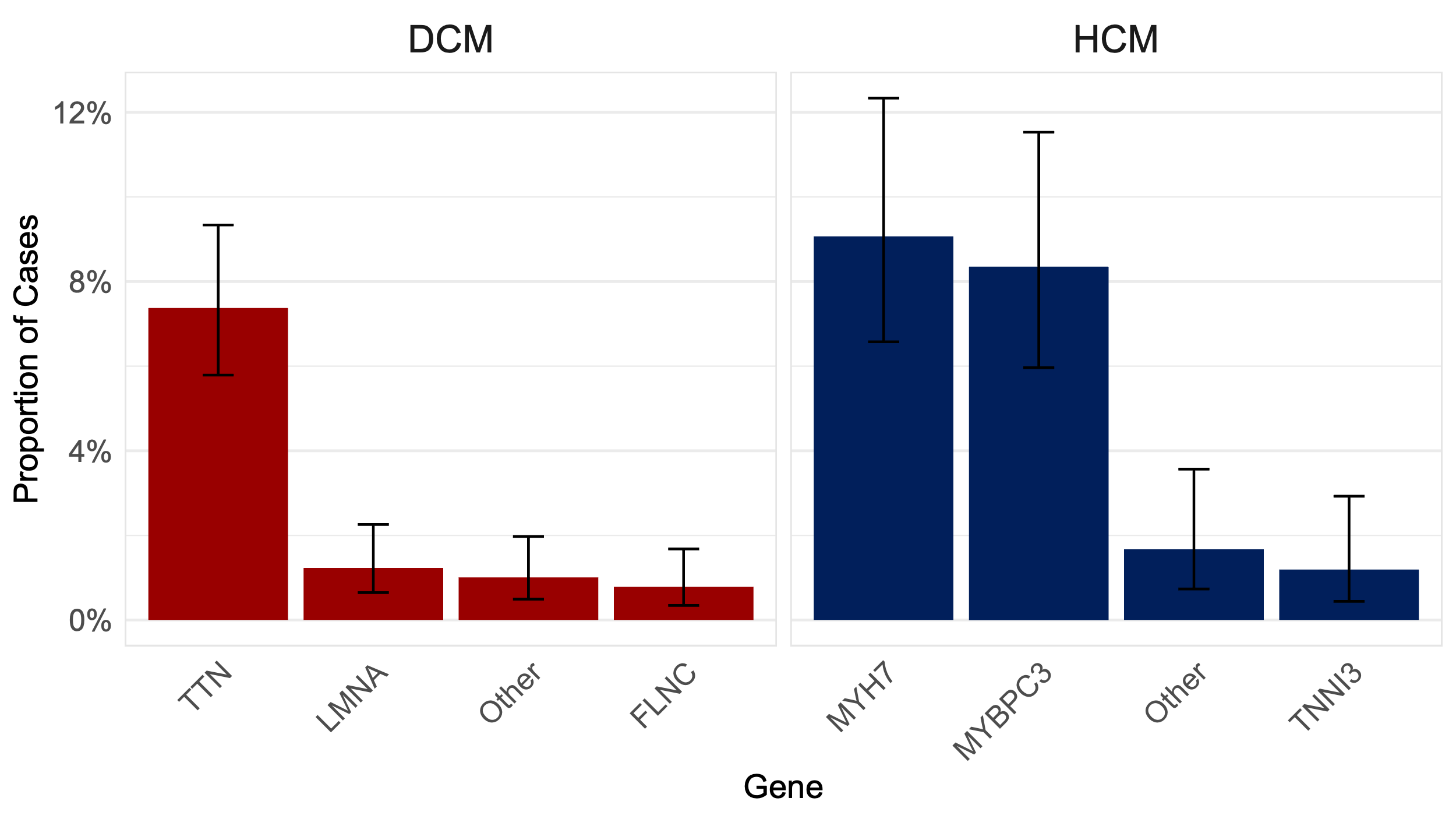

Percentage of DCM (left) and HCM (right) cases with pathogenic variants, by gene. Genes with <5 cases are grouped as “Other.”

#### **eFigure 3: PGS Distributions**

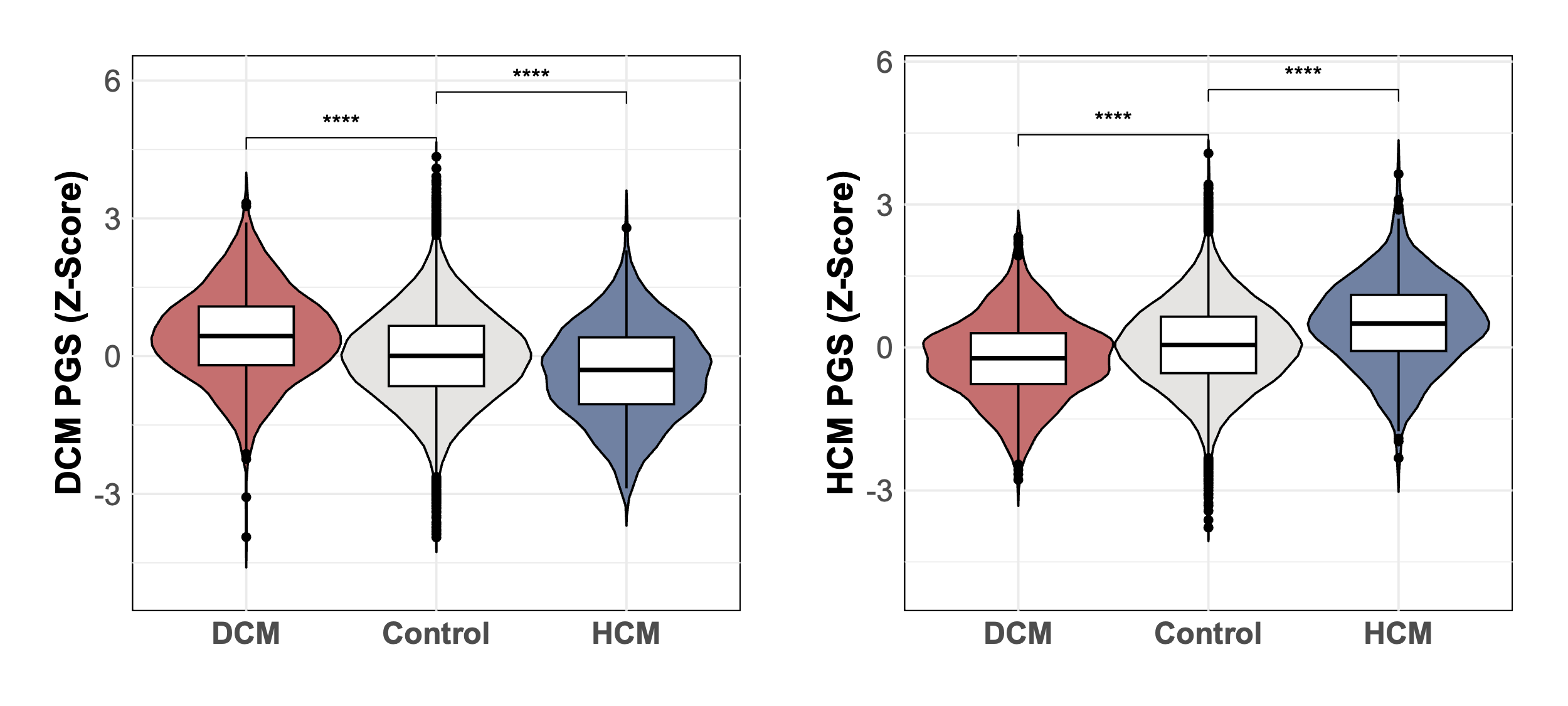

Violin plots of the distribution of DCM PGSs (left panel) and HCM PGSs (right panel) in individuals with DCM (red), HCM (blue), or neither (grey).

#### **eFigure 4: Monogenic and Polygenic Contributions to Calibration of Predictive Models**

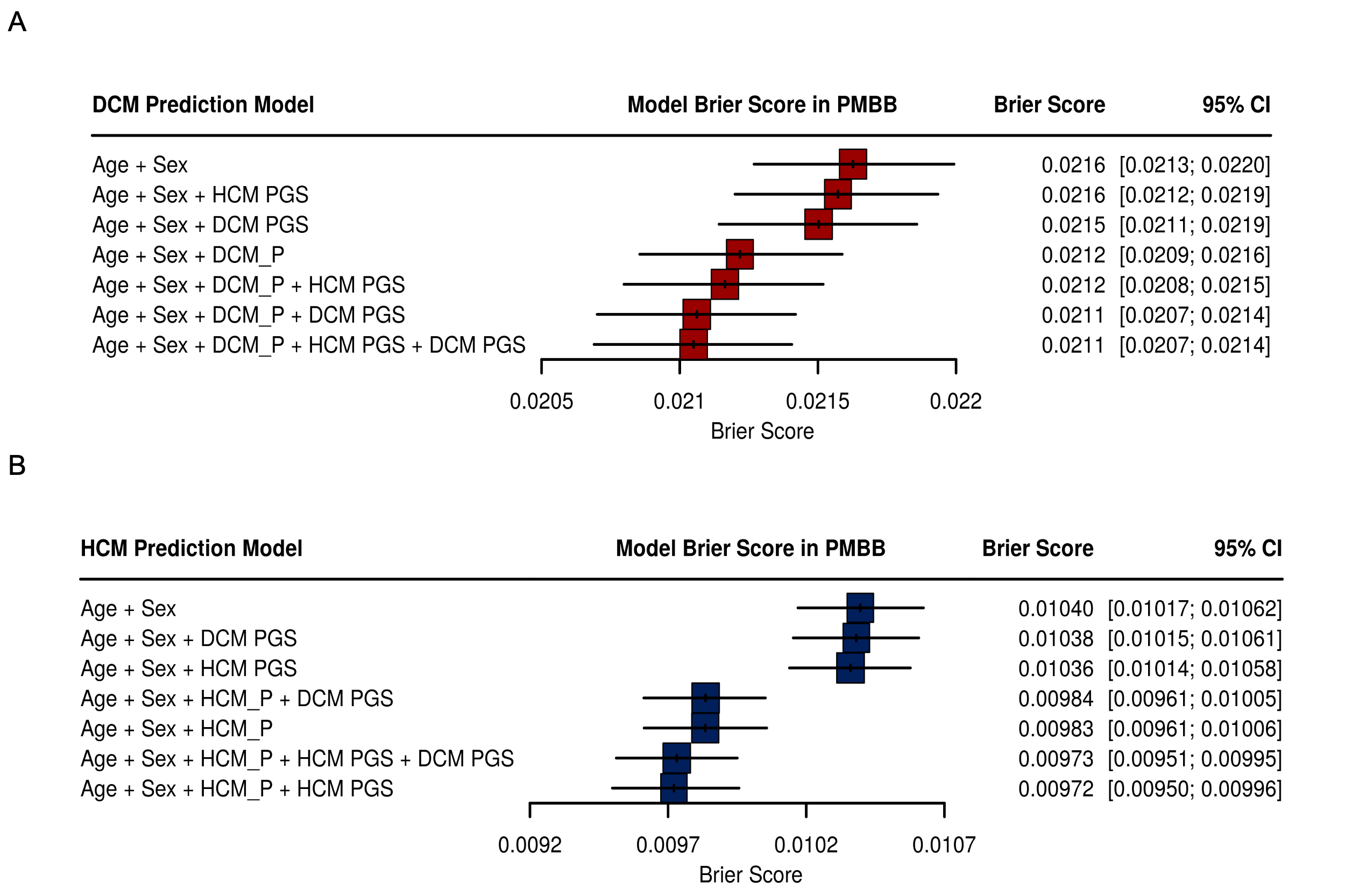
Contributions of monogenic and polygenic risk terms to A) DCM and B) HCM model calibration, measured by Brier Score. HCM_P and DCM_P indicate the presence of a pathogenic variant in an HCM and DCM gene respectively.
